# Widespread cell stress and mitochondrial dysfunction in early Alzheimer’s Disease

**DOI:** 10.1101/2021.08.11.21261851

**Authors:** Ashwin V Venkataraman, Ayla Mansur, Gaia Rizzo, Courtney Bishop, Yvonne Lewis, Ece Kocagoncu, Anne Lingford-Hughes, Mickael Huiban, Jan Passchier, James B Rowe, Hideo Tsukada, David J. Brooks, Laurent Martarello, Robert A. Comley, Laigao Chen, Adam J. Schwarz, Richard Hargreaves, Roger N. Gunn, Eugenii A. Rabiner, Paul M Matthews

## Abstract

Cell stress and impaired oxidative phosphorylation are central to mechanisms of synaptic loss and neurodegeneration in the cellular pathology of Alzheimer’s disease (AD). We quantified the *in vivo* density of the endoplasmic reticulum stress marker, the sigma 1 receptor (S1R) using [^11^C]SA4503 PET, as well as that of mitochondrial complex I (MC1) with [^18^F]BCPP-EF and the pre-synaptic vesicular protein SV2A with [^11^C]UCB-J in 12 patients with early AD and in 16 cognitively normal controls. We integrated these molecular measures with assessments of regional brain volumes and brain perfusion (CBF) measured with MRI arterial spin labelling. 8 AD patients were followed longitudinally to estimate rates of change with disease progression over 12-18 months. The AD patients showed widespread increases in S1R (≤ 27%) and regional decreases in MC1 (≥ -28%), SV2A (≥ -25%), brain volume (≥ -23%), and CBF (≥ -26%). [^18^F]BCPP-EF PET MC1 density (≥ -12%) and brain volumes (≥ -5%) were further reduced at follow up in brain regions consistent with the differences between AD patients and controls at baseline. Exploratory analyses showing associations of MC1, SV2A and S1R density with cognitive changes at baseline and longitudinally with AD, but not in controls, suggested a loss of metabolic functional reserve with disease. Our study thus provides novel *in vivo* evidence for widespread cellular stress and bioenergetic abnormalities in early AD and that they may be clinically meaningful.

## INTRODUCTION

The role of increased oxidative stress and an unfolded protein response (UPR) in early sporadic, late-onset Alzheimer’s Disease (AD) has been highlighted by biochemical and transcriptomic data, as well as genetic and biochemical evidence.*(1)* Multiple factors could contribute to cell stress.

Mitochondrial dysfunction and reduced oxidative phosphorylation have long been recognised in AD.*(2, 3)* Oxidative and ER stress likely are major pathogenic mechanisms in AD. *(4)(5)* The sigma 1 receptor (S1R) is upregulated in the endoplasmic reticulum (ER) at sites of cell stress caused by impaired mitochondrial oxidative phosphorylation. *(6, 7)* Amyloid-β toxicity and impaired autophagy/mitophagy may contribute. These pathologies, as well as inflammatory glial activation, lead to synaptic loss, which has a mechanistically proximate relationship to neuronal circuit dysfunction and cognitive symptoms.*(8)*

However, the long prodromal phase of AD pathology suggests that the healthy brain has considerable functional reserve.*(9)* Functional reserve can be defined operationally as the extent to which redundancy or plasticity allows normal cognition to be maintained despite an impairment of functions (e.g., metabolic, synaptic or neuronal circuit) necessary to support it. Identification of pathological processes affecting pathways without sufficient functional reserve supports prioritisation of those pathways as therapeutic targets for AD, as the correlation of function and cognitive performance suggests that their modulation could lead to measurable clinical benefits.

We sought to characterise relationships between brain cell stress responses, oxidative metabolism and synaptic density in 12 patients with early AD in comparison with 16 healthy controls by imaging with three PET radiotracers to quantify the regional density of relevant molecular effectors. We used [^11^C]SA4503 to image the sigma-1-receptor, [^18^F]BCPP-EF to image mitochondrial complex I, and [^11^C]UCB-J to image the synaptic vesicle glycoprotein 2A (SV2A) protein. Arterial spin labelling (ASL)*(10)* was also performed to quantify brain perfusion along with MRI volumetric assessments. AD patients were followed up 12-18 months later to evaluate the longitudinal change in these markers. By relating these markers of pathology to cognitive performance, we were able to explore bioenergetic and synaptic functional reserve in the AD patients relative to the cognitively normal controls.

## RESULTS

### Patient demographic and neuropsychological results

12 participants had a diagnosis of sporadic onset amnestic, PET amyloid positive early AD and 16 were cognitively normal controls. The two groups had equal numbers of females and males and similar premorbid intelligence levels as assessed by the NART. As expected, the AD patients had a significantly lower MMSE, immediate memory, language and delayed memory test scores compared to the control group but showed no impairment of visuospatial function (Table 1).

**Table 1.** Cross-sectional demographic and neuropsychological testing data for AD patients and cognitively normal controls at baseline.

|  | AD (n=12) |  | CN (n=16) |  | p-value<sup>1</sup> |
| --- | --- | --- | --- | --- | --- |
|  | Mean | SD | Mean | SD |  |
| Age (Years) | 75.50 | 7.82 | 65.88 | 9.82 | **0.008** |
| MMSE | 24.33 | 3.45 | 28.75 | 1.06 | **0.001** |
| Immediate Memory | 79.75 | 23.08 | 105.62 | 11.12 | **0.003** |
| Visuospatial | 101.83 | 17.54 | 98.56 | 9.61 | 0.568 |
| Language | 82.67 | 11.80 | 103.31 | 11.39 | **0.000** |
| Attention | 94.58 | 19.75 | 106.88 | 11.06 | 0.070 |
| Delayed Memory | 65.92 | 26.66 | 102.38 | 10.46 | **0.001** |
| NART | 36.67 | 8.63 | 35.94 | 10.85 | 0.845 |
| <sup>1</sup> Unpaired two-tailed t-test assuming unequal variance |  |  |  |  |  |

Four AD patients did not complete their longitudinal follow-up scans due to progression of AD such that they could not comply with study requirements, new medical illnesses or excessive head motion during scanning. One participant started donepezil, which has high affinity for the S1R, so was excluded from the follow-up [^11^C]SA4503 PET scan. In summary, eight participants with early AD were able to complete both baseline and 12-18 month follow-up SV2A and MC1 PET and neuropsychological testing, and seven participants had both baseline and follow-up S1R PET scans. Ten participants completed baseline and follow up MRI scans. Reductions in ACE memory scores were found in those AD patients assessed longitudinally (Table 2).

**Table 2.**
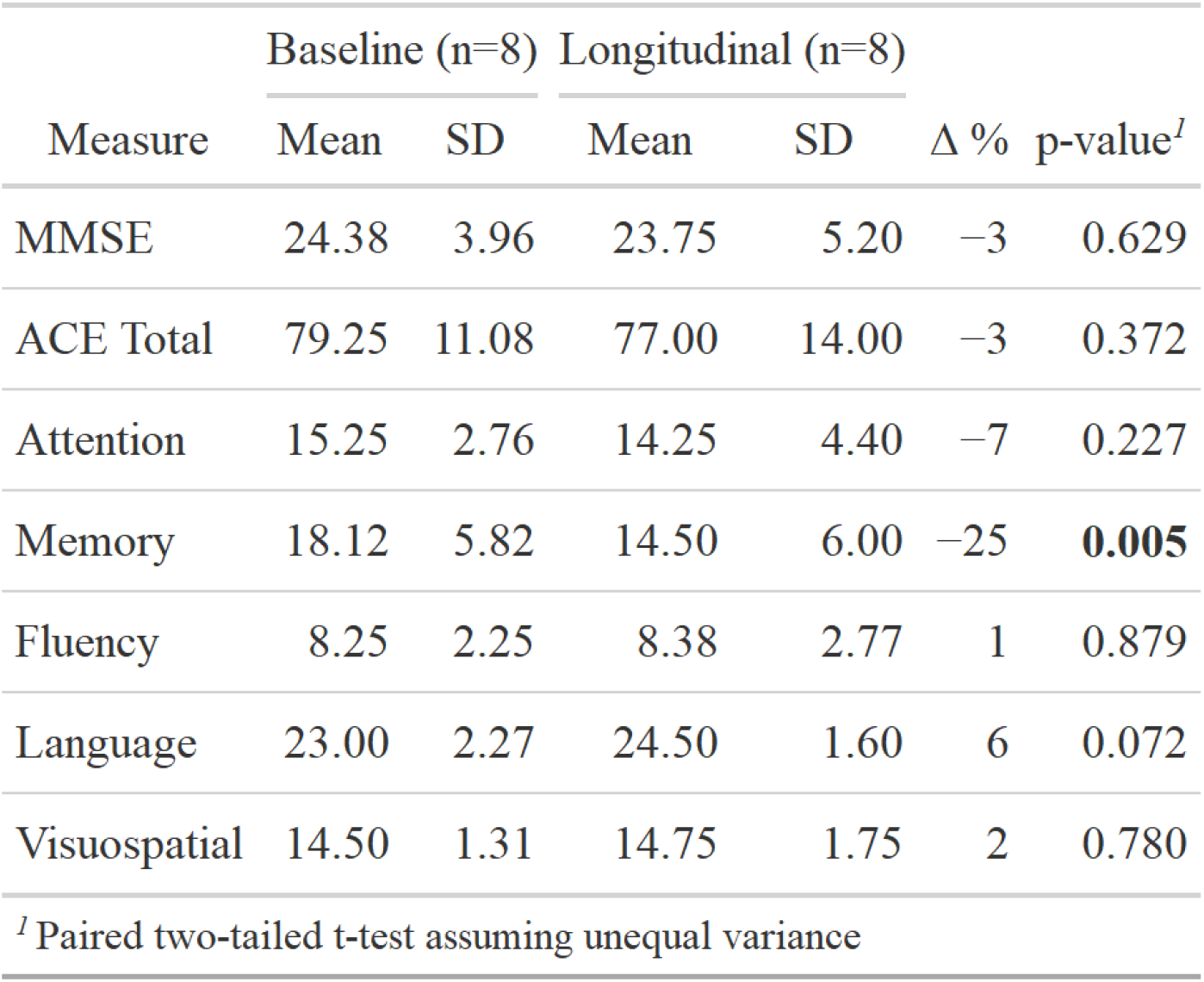
Neuropsychological test results for AD patients after 12-18 months.

### Brain atrophy and reduced brain perfusion in AD brains

At baseline, all ROIs had smaller volumes in the AD patients compared with controls. The greatest relative differences in volumes were in the hippocampus (−23%, p<0.001) and temporal lobe (−16%, p<0.001) (Figure 1A). Partial volume corrected data were used as the primary outcome parameters for the PET and CBF assessments. Uncorrected data was generally consistent with partial volume corrected data and is provided as Supplementary Information.

**Fig. 1.**
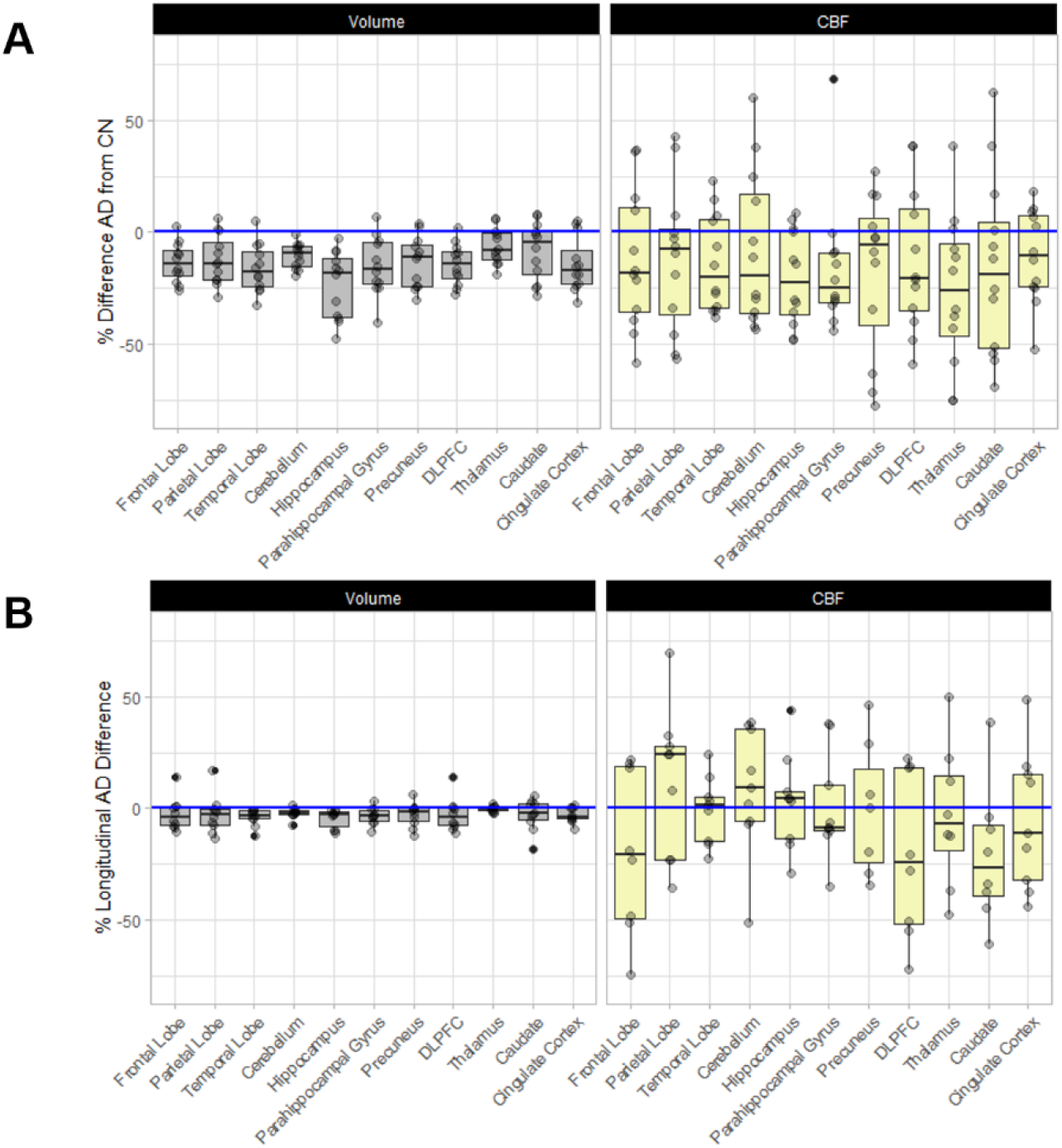
Cross-sectional and longitudinal results for volume and CBF. Figure showing cross-sectional results (A) and longitudinal results (B) for volume (left) and CBF (right). Panel A shows box and whisker plots of individual AD patient percentage differences from the mean controls corresponding with values in Supplementary Table 1. Panel B shows box and whisker plots of longitudinal AD patient percentage change from baseline corresponding with values in Supplementary Table 2

CBF was lower in AD patients compared to controls in all brain regions (range -8 to -26%). The greatest relative reductions in CBF were observed for the hippocampus (−20%, p= 0.023) and thalamus (−26%, p = 0.044) (see Figure 1A).

### Widespread increases in brain oxidative stress responses in AD brains

[^11^C]SA4503 V_T_/f_P_ was increased in all brain ROIs except the hippocampus and caudate in AD patients compared to controls (Figure 2, and 3A). The parametric image contrasting mean voxel-wise [^11^C]SA4503 V_T_/f_P_ for AD patients with the controls was consistent with ROI data (Figure 2) in defining the largest and most statistically significant relative increases in [^11^C]SA4503 binding in the cingulate cortex (+27%, p=0.010), the precuneus (+23%, p= 0.011), cerebellum (+24% p= 0.030) and parietal (+21%, p=0.023) and temporal (+21%, p=0.033) lobes. No significant association was found between [^11^C]SA4503 V_T_/f_P_ and CBF in any of the ROIs for the AD patients or controls.

**Fig. 2.**
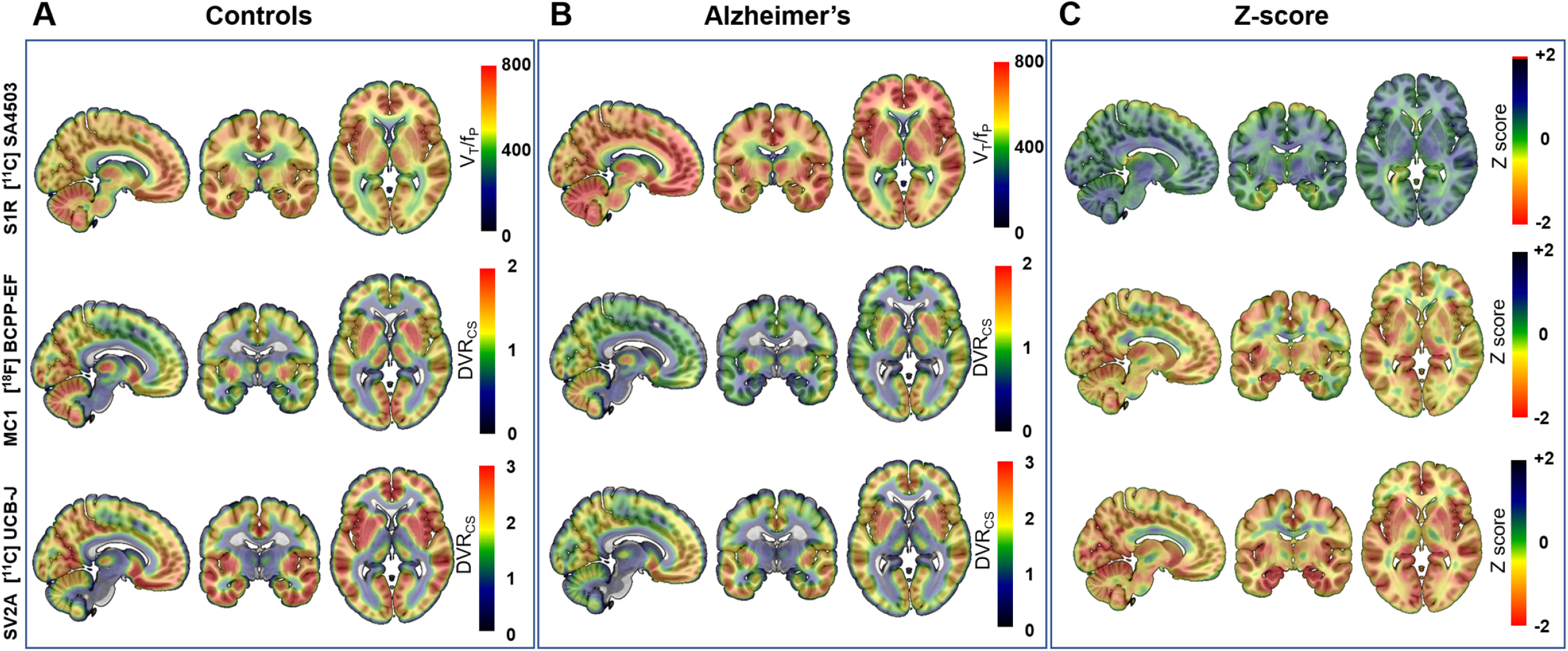
Mean voxel-wise PET images for controls, AD, and z-score contrasting AD from controls. Mean voxel-wise PET images in MNI 152 space for controls (n=16) (A), AD patients at baseline (n=12) (B) and the Z-difference image contrasting the AD patients with controls (C) for [^11^C]SA4503 V_T_/f_P_ (sigma 1 receptor) (top), [^18^F]BCPP-EF DVR_CS_ (mitochondrial complex I) (middle) and [^11^C]UCB-J DVR_CS_ (synaptic density) (bottom).

**Fig. 3.**
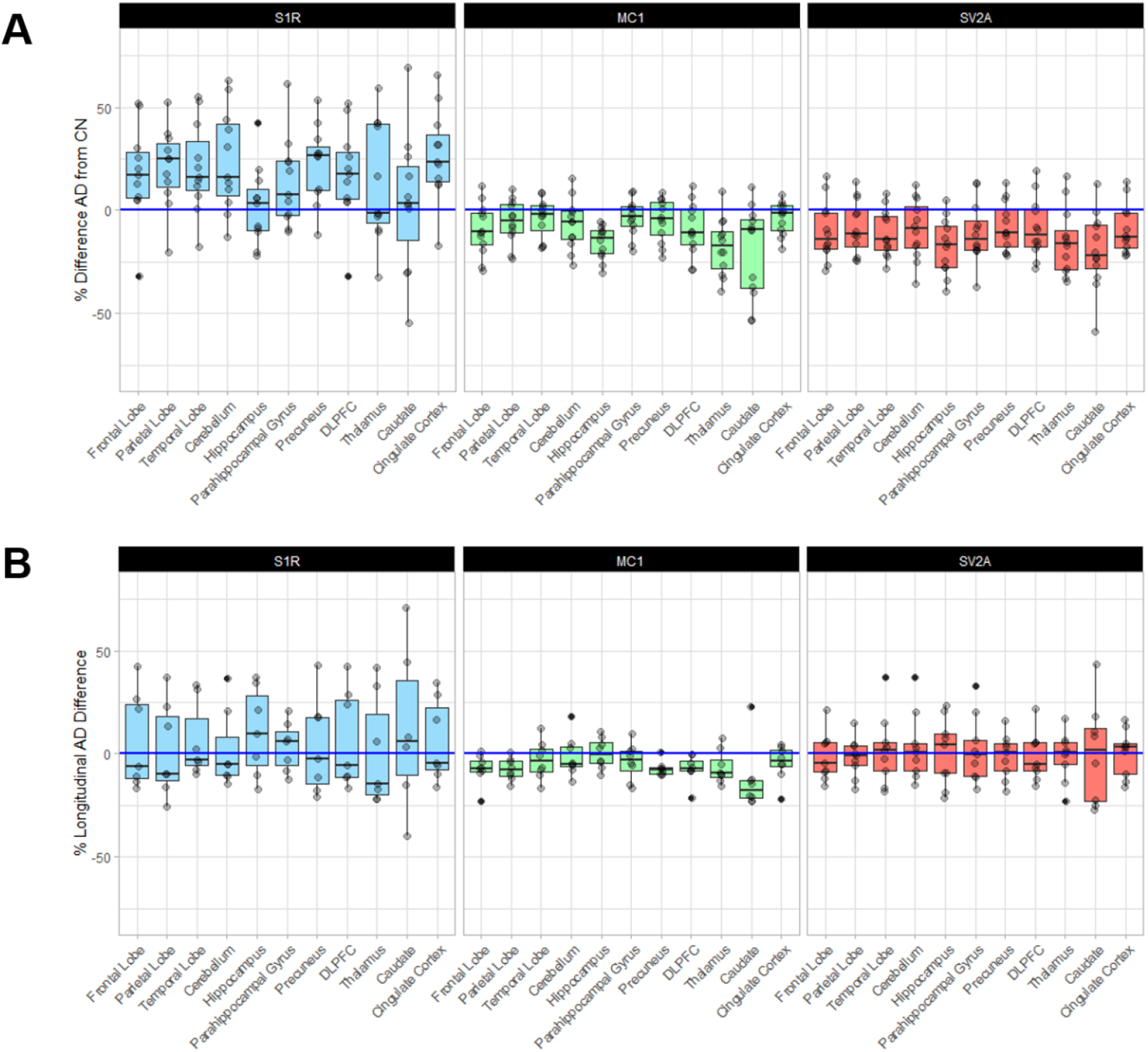
Cross-sectional differences between AD patients and controls, and longitudinal differences for AD patients between S1R, MC1, and SV2A. Cross-sectional differences between regional measures in AD patients (n=12) with the corresponding measures in mean controls (n=16) (A) and changes in individual AD patients between baseline and the 12-18 month follow up (B). Panel A shows box and whisker plots of individual Alzheimer’s patient percentage differences from the mean control values [^11^C]SA4503 V_T_/f_P_ (sigma 1 receptor) (left), [^18^F]BCPP-EF DVR_CS_ (mitochondrial complex I) (middle) and [^11^C]UCB-J DVR_CS_ (synaptic density) (right) (Supplementary Table 1). Panel B shows box and whisker plots of longitudinal AD patient percentage differences at 12-18 month follow up relative to baseline measures (Supplementary Table 2).

### Focal decreases in synaptic density in AD brains

[^11^C]UCB-J DVR_CS_ was generally lower in brains of the AD patients compared with controls, although not significantly reduced in all ROIs (Figure 2 and 3A) The greatest relative reductions in SV2A density were found in the caudate (−25%, p=0.007), hippocampus (−24%, p=0.001) and thalamus (−19%, p=0.012). No significant association was found between [^11^C]UCB-J DVR_CS_ and CBF in any of the ROIs for the AD patients or for controls.

### Focal decreases in mitochondrial complex I in AD brains

[^18^F]BCPP-EF DVR_CS_, which provides a measure of the density of MC1, the first enzyme in the mitochondrial electron transport chain, was lower in the brains of AD patients compared with controls, although it was not significantly reduced in all ROIs (Figure 2, and 3A). The greatest differences in [^18^F]BCPP-EF DVR_CS_ for the AD patients relative to controls were found in the hippocampus (−25%, p = 0.02), thalamus (−23%, p =0.001) and caudate (−28%, p =0.02), with smaller relative reductions in the frontal lobe (−12%, p =0.040) and dorsal lateral prefrontal cortex (DLPFC) (−12%, p =0.048). The parametric images comparing [^18^F]BCPP-EF DVR_CS_between the two groups (Figure 2) highlighted the larger focal differences. We did not find correlations between regional CBF and reduced [^18^F]BCPP-EF DVR_CS_ in ROIs in AD patients or controls.

### Mitochondrial oxidative phosphorylation is uncoupled from synaptic density in AD

MC1 catalyses a first, rate-limiting step in mitochondrial oxidative phosphorylation. [^18^F]BCPP-EF DVR_CS_ and [^11^C]UCB-J DVR_CS_ were positively correlated in controls in all regions, particularly frontal (r^2^=0.67, p=0.0001), parietal (r^2^=0.66, p=0.00012) and hippocampal (r^2^=0.27, p=0.038) cortex, consistent with a physiological coupling between oxidative energy production and synapse density (see Figure 5).*(11)* However, this association was lost in the AD patients (e.g., in the hippocampus r^2^ = 0.058, p=0.45).

### Regional longitudinal changes in metabolic and synaptic markers were consistent with differences between AD patients and controls at baseline

The baseline between group comparisons suggested regional differences in the rates of progression of biochemical and synaptic pathology in AD. We re-scanned the AD group after 12-18 months. As expected, we found small global decreases in brain volume at follow up, with greater relative volume changes in the temporal lobe (−4%, p=0.009), hippocampus (−5%, p=0.01), parahippocampal gyrus (−3%, p=0.027), cingulate cortex (−3%, p=0.023) and cerebellum (−2%, p=0.020). There were no significant changes in CBF (Figure 1B).

Longitudinal PET measures of the biochemical pathology showed regional changes consistent with differences between the AD patients and controls at baseline (Figure 3B)). Regional [^11^C]UCB-J DVR_CS_ was generally lower after 12-18 months, but the changes were small and none were statistically significant. [^11^C]SA4503 V_T_/f_P_ showed non-significant trends for regional increases. By contrast, [^18^F]BCPP-EF DVR_CS_ was decreased in all regions except the hippocampus and was most sensitive to change over time, with the largest relative reductions found in the parietal lobe (−11%, p=0.009), the precuneus (−10%, p=0.001), and the DLPFC (−12%, p=0.037).

Variable increases in [^11^C]SA4503 V_T_/f_P_ with generally greater decreases in [^18^F]BCPP-EF DVR_CS_ and [^11^C]UCB-J DVR_CS_ in the prefrontal cortex were localised in the mean voxel-wise Z-images contrasting follow up with baseline for the AD patients (Figure 4).

**Fig. 4.**
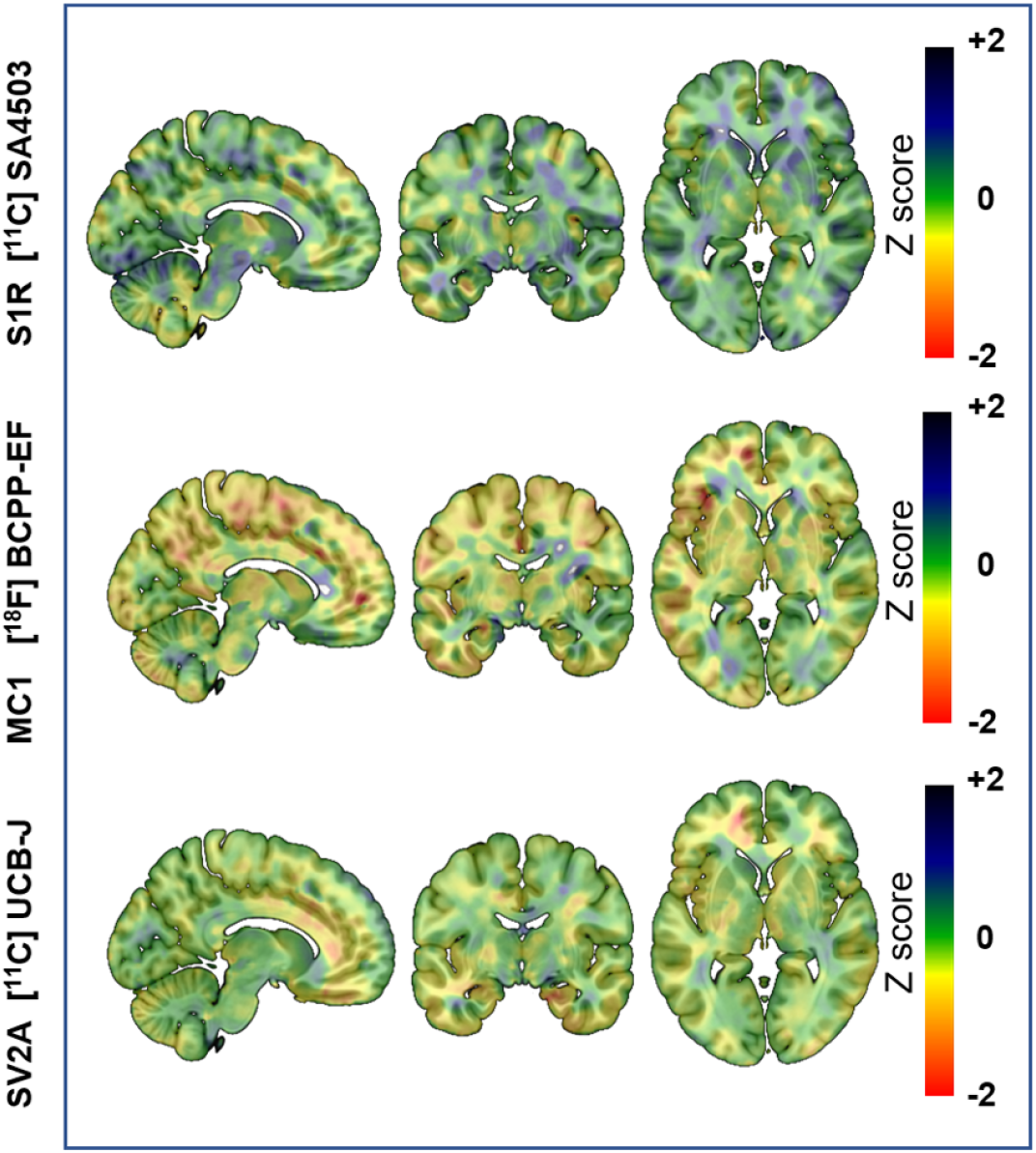
Voxel-wise Z score PET images of longitudinal change in AD patients. Voxel-wise Z score PET images of longitudinal changes in AD patients, contrasting the 12-18 month follow up with baseline images in MNI 152 space for [^11^C]SA4503 V_T_/f_P_ (sigma 1 receptor) (top), [^18^F]BCPP-EF DVR_CS_ (mitochondrial complex I) (middle) and [^11^C]UCB-J DVR_CS_ (synaptic density) (bottom).

**Fig. 5.**
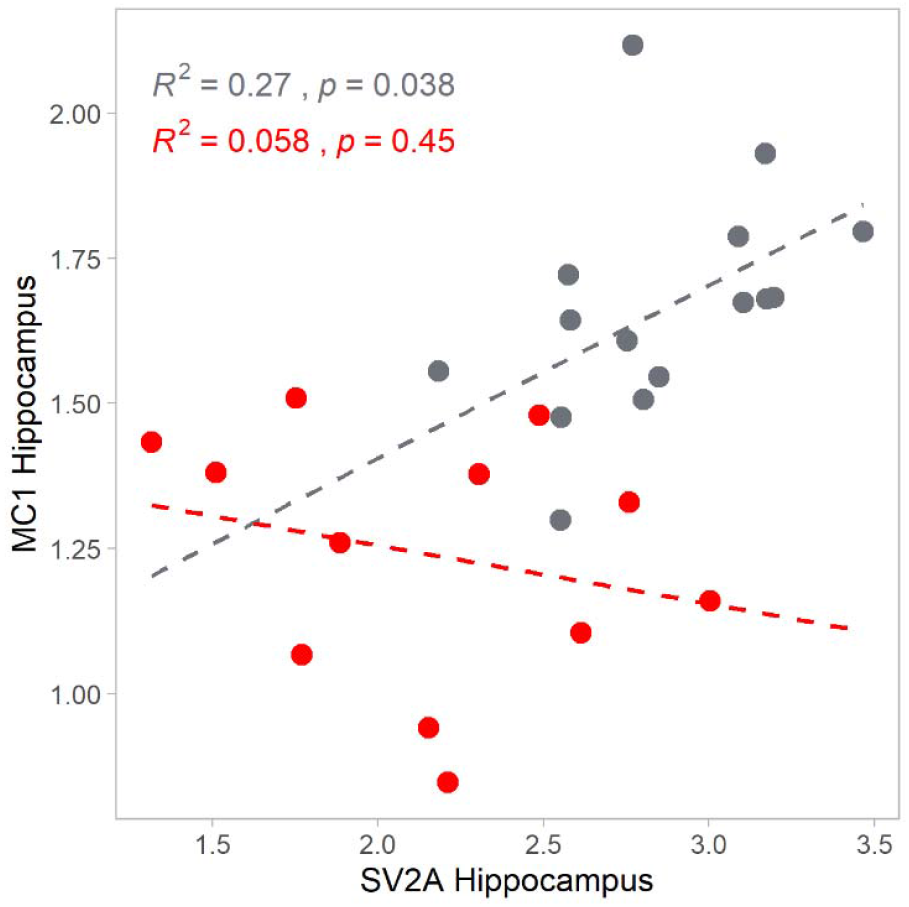
Associations between MC1 and SV2A in the hippocampus for controls and AD patients. Associations between [^18^F]BCPP-EF DVR_CS_ (MC1, ordinate) and [^11^C]UCB-J DVR_C_ (SV2A, abscissa) in hippocampus for controls (grey) and AD patients (red). [^18^F]BCPP-EF DVR_CS_ and [^11^C]UCB-J DVR_CS_ were moderately to strongly positively correlated in controls all regions, consistent with a physiological coupling between oxidative energy production and synapse density,*(11)* and this association was reduced or absent with Alzheimer’s disease.

### Loss of functional reserve in AD

We explored whether the AD patients had reduced brain functional reserve by exploring correlations between measures of mitochondrial oxidative capacity, synaptic density, and cell stress and measures of cognitive performance. [^18^F]BCPP-EF DVR_CS_ showed negative associations with visuospatial scores in the AD patients (r^2^ = 0.45, p=0.016), but not in controls (r^2^=0.076, p=0.3). Hippocampal [^11^C]SA4503 V_T_/f_P_ showed significant negative associations with visuospatial scores in AD patients (r^2^ = 0.57, p=0.0076) not found for the controls (r^2^ = 0.0061, p=0.77). Associations of cognitive performance with synaptic density [^11^C]UCB-J DVR_CS_ or language function tests (picture naming and semantic fluency) showed strong positive correlations in the AD patients (hippocampus, r^2^ = 0.68, p=0.0001, parietal lobe, r^2^ = 0.39, p=0.03), but not in controls (hippocampus, r^2^ = 0.0004, p=0.94, parietal lobe, r^2^ = 0.009, p = 0.73). Finally, we explored correlates of changes in memory over time in the AD patients. Longitudinal *decreases* in frontal [^18^F]BCPP-EF DVR_CS_ (r^2^ = 0.61, p=0.021) were associated with decreases in ACE memory scores. Longitudinal *increases in* frontal lobe [^11^C]SA4503 V_T_/f_P_ were associated with decreases in ACE memory scores (r^2^ = 0.64, p=0.031).

## DISCUSSION

Our study is unique for its *in vivo* multi-modal, longitudinal characterisation of molecular markers related to mitochondrial and synaptic dysfunction in early AD. We exploited recently introduced PET markers of cellular stress, mitochondrial and synaptic density to explore *in vivo* pathology in AD. Highly specific molecular markers were complemented by more generalised physiological perfusion (ASL) and volumetric MRI measures. Observation of increased S1R density in AD provides evidence of a generalised ER stress response. More focal decreases in synaptic and mitochondrial density, seen particularly in the hippocampus, thalamus and caudate, suggest selective regional vulnerability to this generalised biochemical stress. The positive correlation between hippocampal mitochondrial and synaptic density found in controls was not seen in the AD patients, providing evidence for a loss of the normal physiological relationship between mitochondrial oxidative phosphorylation and synaptic density in disease.*(11)* Longitudinal follow-up of the AD patients showed that the measures of mitochondrial activity and brain volume loss were most sensitive to disease progression, although non-significant changes in other measures were broadly consistent with their relative differences between AD and controls at baseline. Exploratory analyses showed associations between regional mitochondrial, synaptic and sigma-1 receptor density (a marker of cellular stress) with impaired visuospatial function, language and memory test performance in the AD patients, but not in controls, suggesting loss of synaptic and metabolic functional reserve with disease. Overall, our results are consistent with a disease model in which generalised cell stress leads to regionally variably expressed pathologies of mitochondrial dysfunction, synaptic loss and clinical expression of AD.

The S1R is a pluripotent modulator of multiple systems*(12)* found at the ER-mitochondrial interface (MAM) and expressed widely throughout the CNS*(13)*. Increased expression of S1R provides an index of adaptive responses to cell stress and [^11^C]SA4503 PET imaging has been validated as a measure of the distribution of S1R in the brain*(14) (15) (16)*. Using [^11^C]SA4503 PET imaging, we have provided evidence for the first time of widespread and generalised increases in S1R density in early AD patients. An earlier study reported that S1R binding was *decreased* in AD*(17)*, but the investigators scanned participants who were being treated with donepezil, which confounds interpretation as it has high affinity for S1R*(18)* and thus blocks the binding of [^11^C]SA4503 to the S1R. We hypothesise that the generalized increase in S1R binding in AD patients is a consequence of interactions between aging and genetic risk susceptibility that act throughout the brain. However, our observation that S1R binding was not increased in patients with AD in the hippocampus, the brain region that showed the biggest differences in mitochondrial and synaptic density compared to controls, suggests that expression of S1R also must be influenced by regionally specific factors.

MC1 is the first enzyme in the mitochondrial electron transport chain.*(19)* [^18^F]BCPP-EF was developed as a PET radioligand to image MC1 density *in vivo(20–22)* and the first full human characterisation was completed only recently*(16, 23)*. We found decreases in MC1 binding in AD patients greater than previously reported*(24, 25)*, most prominently in the hippocampus and in the highly connected subcortical grey matter of the caudate and thalamus. [^18^F]BCPP-EF DVR_CS_ was the most sensitive longitudinal marker of disease progression of those that we explored. The observation that CBF and MC1 binding were not correlated in AD patients argues that the impaired mitochondrial biogenesis or dysfunction is independent of perfusion deficits.*(26)*

Synaptic loss begins early and progresses in AD*(27). In vivo* human imaging studies based on PET radioligand binding to the synaptic vesicle glycoprotein 2A (SV2A)*(28, 29)* showing reduced tissue SV2A density highlight that relative cortical volume reductions*(30)* underestimate the extent of synaptic loss in AD. However, although we observed some volume loss over the short 12-18 month longitudinal follow up, we could not measure significant reductions of SV2A density in the remaining brain tissue. Future studies with larger numbers of subjects or longer periods of follow up will be needed to estimate the rate of SV2A marker loss independent of the progressive brain atrophy in AD and to identify where and when it is most sensitive to disease progression.

As well as underestimating the magnitude of synaptic loss across the brain, its distribution also is not fully explained by the regional brain volume loss. We found large reductions in SV2A in AD patients grey matter, particularly in the hippocampus, caudate and thalamus. As there is less relative atrophy of the subcortical nuclei in AD disease than in the hippocampus, we speculate that the high synaptic loss seen in these nuclei reflects loss of convergent projections from widespread cortical regions, in addition to any local neurodegeneration.

Cognitively normal controls did not show meaningful associations of cognitive performance measures with levels of S1R, MC1 or SV2A, consistent with functional reserve in the healthy older brain. Visuospatial, language and memory performance were correlated with hippocampal and frontal lobe S1R, MC1 or SV2A for AD patients. This suggests that functional reserve present in the healthy brain is lost in early AD, in which mitochondrial activity, oxidative stress responses and synaptic density appear to become rate limiting for cognition. If these correlations can be shown to reflect causal relations, therapeutic targeting of the responsible mechanisms could provide clinical benefits and be monitored using the measures here.

Although this study had a relatively small group of AD patients and short period of follow up, our study illustrates the potential value of multi-modal, longitudinal imaging studies in AD, where no single measure describes a sufficient breadth of pathology to fully characterise disease progression. Ours is the first such study to date to include longitudinal observations which support and extend cross-sectional ones. The results emphasise that, despite the magnitude of regional changes (e.g., in the hippocampus), cell stress responses in AD are generalised, suggesting a widespread, prodromal biochemical pathology. Elucidating mechanisms linking genetic susceptibility to this biochemical pathology will be important for prioritising targets for earliest interventions to delay or reverse disease.

## MATERIALS AND METHODS

### Study Design

We included 28 participants. Twelve patients with amnestic early AD were recruited for this study, and 16 controls were drawn from the MINDMAPS healthy volunteers cohort. Molecular Imaging of Neurodegeneration – Mitochondria, Associated Proteins, Synapses (MINDMAPS) is an academic/industry consortium to investigate neurodegenerative diseases using these PET targets (invicro.com/mind-maps). All participants underwent screening, [^18^F]BCPP-EF, [^11^C]UCB-J, [^11^C]SA4503 dynamic PET imaging, volumetric MRI and ASL scanning. All MR and PET measures were repeated in 8 of the AD patients 12-18 months after the baseline imaging.

AD patients met National Institute on Aging-Alzheimer’s Association (NIA-AA) core clinical criteria for probable AD dementia, or amnestic mild cognitive impairment. They were beta-amyloid positive (based on [^18^F]Florbetaben PET findings), were aged >50, had a Mini Mental State Examination (MMSE) score ≥18, and had the capacity to give informed consent. Participants taking symptomatic therapy for AD were on a stable dose for at least 6 weeks prior to the baseline evaluation, and were willing to adhere to PET scan procedures.

Exclusion criteria for both groups included major psychiatric/neurological/ medical illness or infection, use of medication (e.g. donepezil) known to either bind directly to SV2A, S1R or MC1, or impair cognition, and prior radiation exposure >10mSv in the past year. Routine clinical blood samples alongside ApoE genotyping were performed.

Neuropsychological and cognitive assessments included the Mini Mental State Examination (MMSE), Addenbrooke’s Cognitive Exam (ACE-III), Repeatable Battery for Assessment of Neuropsychological Status (RBANS) and the National Adult Reading Test (NART). Specific RBANS tests included assessing immediate memory (list learning and story memory), visuospatial perception (figure copy and line orientation), language (picture naming and semantic fluency), attention (digit span and coding), and delayed memory (list recall, list recognition, story memory and figure recall). Specific ACE-III tests included rating attention (orientation), memory (word recall, address recall, semantic recall), fluency (word generation), language (sequenced commands, writing, repetition, pronunciation) and visuospatial (figure copy, clock drawing, dot counting, letter recognition).

All participants provided written informed consent. Screening and scanning were conducted at the Invicro imaging centre, Hammersmith Hospital Site, London. Ethical approval for this study was provided by the NHS London - Brighton & Sussex Research Ethics Committee (REC 18/LO/0179) for AD patients and East of England Cambridge Central & South Research Ethics Committee for controls. Radiation safety was approved by the Administration of Radioactive Substances Advisory Committee (ARSAC R92). Local site approval was provided by Imperial College London Joint Research Office. Participants were recruited from Imperial College Healthcare NHS Trust, West London Mental Health Trust, and Cambridge University Hospitals.

### Image Acquisition

[^18^F]BCPP-EF, [^11^C]UCB-J, and [^11^C]SA4503 were synthesised and PET/CT images and arterial blood samples acquired as previously described*(16, 20, 28, 31)*. [^18^F]Florbetaben was purchased from Alliance Medical. Imaging was performed using either a Siemens Biograph 6 True Point or HiRez PET/CT scanner (Siemens Healthcare, Erlangen, Germany). Prior to each imaging session a low-dose CT scan was acquired to enable attenuation correction (30 mAs, 130 KeV, 0.55 pitch). AD patients received a [^18^F]Florbetaben scan at screening, acquired over 20 min following a 90 min distribution phase. All participants received [^18^F]BCPP-EF, [^11^C]UCB-J and [^11^C]SA4503 scans with the same camera which was also used for all follow-up scans for an individual. The PET tracers were administered as an intravenous bolus (20mls over 20 seconds), and blood samples were collected from the radial artery to assess changes in the concentration of radioactivity in whole blood and plasma, as well as the percent of unmetabolized tracer over the course of the PET scan radioactivity sample collection. Following tracer administration, dynamic emission data was acquired in list mode over 90 minutes and reconstructed into 26 time frames (frame durations: 8×15 s, 3×60 s, 5×120 s, 5×300 s, 5×600 s) using discrete inverse Fourier transform reconstruction with corrections for attenuation, randoms and scatter.

All MRIs were acquired with a Siemens 3T Trio clinical scanner (Siemens Healthineers, Erlangen, Germany) using a 32-channel phased-array head coil. T1-weighted structural data (a 3D MPRAGE sequence; TE = 2.98 ms, TR = 2300 ms, flip angle = 9°, voxel size = 1.0 mm x 1.0 mm x 1.0 mm) and multi-TI Pulsed ASL (5 TIs: 1200-2400ms with 300ms increments) are presented in this paper. All measures were repeated 12-18 months later for the AD group.

### Image Analysis and statistical analysis

[^18^F]Florbetaben data were quantified by Amyloid IQ*(32)*, using 33% as cut-off for amyloid positivity and inclusion in the study.

The dynamic PET image analysis pipeline, including generation of an arterial input function, tracer kinetic modelling, model comparison and selection and time stability assessment, has been described previously*(16)*. Briefly, PET scan emission data were processed using MIAKAT™ (version 4.3.24, miakat.org). MIAKAT™ is implemented using MATLAB (version R2016a; Mathworks) and makes use of SPM12 (Wellcome Trust Centre for Neuroimaging, fil.ion.ucl.ac.uk/spm) functions for image segmentation and registration. Individual participant structural MRIs underwent grey matter segmentation and rigid body co-registration to a standard reference space*(12)*. The template brain image and associated CIC neuroanatomical atlas was then nonlinearly warped to the individual participant’s MRI where the regions of interest (ROI) were defined and volumes of regions in mm^3^ were measured. A centrum semiovale (CS) ROI was also generated from the automated anatomic labelling template as defined previously for use as a non-specific reference region for [^11^C]UCB-J.*(28)* PET images were registered to each participant’s MRI and corrected for motion using frame-to-frame rigid-body registration. Regional time activity curves (TAC) were generated for each ROI.

For ASL processing, T1w data was initially processed using FSL (*fsl_anat*) to provide tissue segmentations and nonlinear mapping to standard MNI 152 space (fsl.fmrib.ox.ac.uk)*(33)*. By inversion of this nonlinear transformation, and concatenation with the T1-to-ASL (M0) alignment using FSL (flirt, 6 DOF), the CIC atlas was nonlinearly aligned to the ASL data to provide native-space ROIs. After motion-correction of the ASL data using FSL’s *mcflirt,(34)* and generation of the perfusion-weighted time-series, CBF and arterial arrival time (AAT) modelling was performed using FSL’s *oxford_asl* tool.

Regional volumes of distribution (V_T_) were calculated for each of the three radioligands using the one-tissue compartmental model (1TC) for [^11^C]UCB-J, and the multilinear analysis 1 (MA1) model for [^18^F]BCPP-EF and [^11^C]SA4503 as described previously*(16)*. The outcome parameter chosen for [^11^C]UCB-J and [^18^F]BCPP-EF was the DVR_CS_, with both using the CS as a pseudo-reference region where

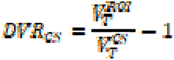

with 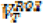 being the V_T_ in the region of interest and 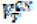 being the V_T_ in the centrum semiovale, a white matter pseudo-reference region. The volume of distribution corrected for plasma fraction (VT/f_p_) was chosen as the outcome parameter for [^11^C]SA4503. Partial volume-corrected data were generated using the Muller-Gartner algorithm.*(35)*

Parametric images in MNI152 space were expressed as z-scores to facilitate interpretation of findings across the three radiotracers. [^11^C]SA4503 and [^11^C]UCB-J parametric maps were generated with the one tissue reversible compartmental model, solved voxel-wise using the basis function method.*(36)* [^18^F]BCPP-EF parametric maps were generated using the MA1 method solved at the voxel level, using the same settings as the region level analysis. Group cross-sectional data of AD patients compared to controls was computed as z-scores (AD patients mean – controls mean)/(controls standard deviation). Group longitudinal compared with baseline AD patient data were also computed as z-scores (AD patients longitudinal mean – AD patients baseline mean)/(AD patients baseline standard deviation).

Comparison of regional target binding between patient and control groups was performed using a two-tailed, unpaired Student’s t-test for cross-sectional data and a two-tailed, paired Student’s t-test for longitudinal data across *a priori* selected regions (frontal lobe, parietal lobe, temporal lobe, cerebellum, hippocampus, parahippocampal gyrus, precuneus, dorsolateral prefrontal cortex, thalamus, caudate, and cingulate cortex). We did not correct for multiple comparisons owing to the small sample size and exploratory nature of this study; however, adjusted p-values using the False Discovery Rate are documented in the supplementary material.

Pearson’s r correlations were performed across modalities and groups for [^11^C]UCB-J, [^18^F]BCPP-EF, and [^11^C]SA4503 uptake, regional volumes and CBF. Associations between imaging findings, neuropsychological and cognitive performance variables were interrogated. Correlations of differences over time and neuropsychological and cognitive performance measures were also explored for *a priori* hypotheses. All statistics were analysed and plotted using R software (R-project.org).

## Data Availability

The data for this study is managed within the MINDMAPS consortium. Information concerning access is available from the lead

## Acknowledgments

We would like to thank all participants and their carers, patient and public involvement (PPI) groups and the Alzheimer’s Society Research Network Monitors who helped with this study.

We would like to acknowledge the excellent and invaluable technical and administrative support provided by the Invicro-London radiochemistry, medical physics, blood laboratory, medical, nursing, PET technologist and radiography teams, to enable the collection of our data.

A.V.V. is supported by the Alzheimer’s Society, grant number 440 (AS-CTF-18-006) and has received support from the NIHR Biomedical Research Centre at Imperial College London.

P.M.M. acknowledges generous personal and research support from the Edmond J Safra Foundation and Lily Safra, an NIHR Senior Investigator Award, the UK Dementia Research Institute and the NIHR Biomedical Research Centre at Imperial College London.

J.B.R. is supported by the NIHR Cambridge Biomedical Research Centre (BRC-1215-20014), and Medical Research Council (SUAG/051 G101400; MR/L023784/2; MR/N029941/1).

This work is supported by the UK Dementia Research Institute which receives its funding from UK DRI Ltd, funded by the UK Medical Research Council, Alzheimer’s Society and Alzheimer’s Research UK. The views expressed are those of the authors and not necessarily those of the NIHR or the Department of Health and Social Care.

## Author contributions

Conceptualization: AVV, ALH, MH, JP, JBR, HT, DJB, LM, RAC, LC, AJS, RH, RNG, EAR and PMM

Methodology: AM, GR, CB, RNG, EAR and PMM

Investigation: AVV

Study regulation, recruitment: AVV, YL, EK

Formal Analysis: AVV

Visualization: AVV

Funding acquisition: EAR, PMM

Supervision: EAR, PMM

Writing – original draft: AVV, EAR, PMM

Writing – review & editing: All authors

## Competing interests

AVV and ALH declare that they have no competing interests.

PMM acknowledges consultancy fees from Novartis, Bristol Myers Squibb, Celgene and Biogen He has received honoraria or speakers’ honoraria from Novartis, Biogen and Roche and has received research or educational funds from Biogen, Novartis, GlaxoSmithKline and Nodthera.

AM, GR, CB, YL, MH, JP, RNG, EAR are employees of Invicro, a Konica Minolta Company.

JBR serves as Editor to Brain; Chief Scientific Advisor to ARUK; and have consultancies with

UCB, Asceneuron, Biogen, WAVE, SV Health, Astex unrelated to the current work; and have research grants from Janssen, Lilly, AstraZeneca.

LM is an employee of Biogen.

RC is an employee and shareholder of AbbVie, Inc.

LC is an employee and shareholder of Pfizer, Inc.

AJS is a full-time employee and shareholder of Takeda Pharmaceuticals, Ltd.

HT is an employee of Hamamatsu Photonics The patent of JP 2014-531670 for

[^18^F]BCPP-EF has been filed by Hamamatsu Photonics KK.

RH is a full-time employee and shareholder of Bristol Myers Squibb.

## REFERENCES AND NOTES

1. E. Tönnies, E. Trushina, Oxidative Stress, Synaptic Dysfunction, and Alzheimer’s Disease, J. Alzheimer’s Dis. 57, 1105–1121 (2017).

2. R. H. Swerdlow, Mitochondria and Mitochondrial Cascades in Alzheimer’s Disease, J. Alzheimer’s Dis. 62, 1403–1416 (2018).

3. D. A. Butterfield, B. Halliwell, Oxidative stress, dysfunctional glucose metabolism and Alzheimer disease, Nat. Rev. Neurosci. 20, 148–160 (2019).

4. W. J. Huang, X. Zhang, W. W. Chen, Role of oxidative stress in Alzheimer’s disease (review), Biomed. Reports 4, 519–522 (2016).

5. S. Hashimoto, T. C. Saido, Critical review: Involvement of endoplasmic reticulum stress in the aetiology of Alzheimer’s disease, Open Biol. 8 (2018), doi:10.1098/rsob.180024.

6. J. Wang, A. Shanmugam, S. Markand, E. Zorrilla, V. Ganapathy, S. B. Smith, Sigma 1 receptor regulates the oxidative stress response in primary retinal Müller glial cells via NRF2 signaling and system xc(-), the Na(+)-independent glutamate-cystine exchanger., Free Radic. Biol. Med. 86, 25–36 (2015).

7. B. Penke, L. Fulop, M. Szucs, E. Frecska, The Role of Sigma-1 Receptor, an Intracellular Chaperone in Neurodegenerative Diseases, Curr. Neuropharmacol. 16, 97–116 (2017).

8. L. Rajendran, R. C. Paolicelli, Microglia-mediated synapse loss in Alzheimer’s disease, J. Neurosci. 38, 2911–2919 (2018).

9. Y. Stern, Cognitive reserve in ageing and Alzheimer’s disease, Lancet Neurol. 11, 1006–1012 (2012).

10. P. Jezzard, M. A. Chappell, T. W. Okell, Arterial spin labeling for the measurement of cerebral perfusion and angiography, J. Cereb. Blood Flow Metab. 38, 603–626 (2017).

11. J. J. Harris, R. Jolivet, D. Attwell, Synaptic Energy Use and Supply, Neuron 75, 762–777 (2012).

12. T.-P. Su, T.-C. Su, Y. Nakamura, S.-Y. Tsai, The Sigma-1 Receptor as a Pluripotent Modulator in Living Systems, Trends Pharmacol. Sci. 37, 262–278 (2016).

13. R. R. Matsumoto, in Sigma Receptors, (Springer US, Boston, MA), pp. 1–23.

14. K. Matsuno, M. Nakazawa, K. Okamoto, Y. Kawashima, S. Mita, Binding properties of SA4503, a novel and selective σ1 receptor agonist, Eur. J. Pharmacol. 306, 271–279 (1996).

15. M. Sakata, Y. Kimura, M. Naganawa, K. Oda, K. Ishii, K. Chihara, K. Ishiwata, Mapping of human cerebral sigma1 receptors using positron emission tomography and [11C]SA4503, Neuroimage 35, 1–8 (2007).

16. A. Mansur, E. A. Rabiner, R. A. Comley, Y. Lewis, L. T. Middleton, M. Huiban, J. Passchier, H. Tsukada, R. N. Gunn, Characterization of 3 PET Tracers for Quantification of Mitochondrial and Synaptic Function in Healthy Human Brain: 18 F-BCPP-EF, 11 C-SA-4503, and 11 C-UCB-J, J. Nucl. Med. 61, 96–103 (2020).

17. M. Mishina, M. Ohyama, K. Ishii, S. Kitamura, Y. Kimura, K. I. Oda, K. Kawamura, T. Sasaki, S. Kobayashi, Y. Katayama, K. Ishiwata, Low density of sigma1 receptors in early Alzheimer’s disease, Ann. Nucl. Med. 22, 151–156 (2008).

18. N. K. Ramakrishnan, A. K. D. Visser, M. Schepers, G. Luurtsema, C. J. Nyakas, P. H. Elsinga, K. Ishiwata, R. A. J. O. Dierckx, A. Van Waarde, Dose-dependent sigma-1 receptor occupancy by donepezil in rat brain can be assessed with 11C-SA4503 and microPET, Psychopharmacology (Berl). 231, 3997–4006 (2014).

19. L. A. Sazanov, A giant molecular proton pump: Structure and mechanism of respiratory complex I, Nat. Rev. Mol. Cell Biol. 16, 375–388 (2015).

20. N. Harada, S. Nishiyama, M. Kanazawa, H. Tsukada, Development of novel PET probes, [18F]BCPP-EF, [ 18F]BCPP-BF, and [11C]BCPP-EM for mitochondrial complex 1 imaging in the living brain, J. Label. Compd. Radiopharm. 56, 553–561 (2013).

21. S. Kazami, S. Nishiyama, Y. Kimura, H. Itoh, H. Tsukada, BCPP compounds, PET probes for early therapeutic evaluations, specifically bind to mitochondrial complex I, Mitochondrion 46, 97–102 (2019).

22. H. Tsukada, H. Ohba, M. Kanazawa, T. Kakiuchi, N. Harada, Evaluation of 18F-BCPP-EF for mitochondrial complex 1 imaging in the brain of conscious monkeys using PET, Eur. J. Nucl. Med. Mol. Imaging 41, 755–763 (2014).

23. A. Mansur, E. A. Rabiner, H. Tsukada, R. A. Comley, Y. Lewis, M. Huiban, J. Passchier, R. N. Gunn, Test–retest variability and reference region-based quantification of 18 F-BCPP-EF for imaging mitochondrial complex I in the human brain, J. Cereb. Blood Flow Metab. 41, 771–779 (2021).

24. T. Terada, T. Obi, T. Bunai, T. Matsudaira, E. Yoshikawa, I. Ando, M. Futatsubashi, H. Tsukada, Y. Ouchi, In vivo mitochondrial and glycolytic impairments in patients with Alzheimer disease, Neurology 94, e1592–e1604 (2020).

25. T. Terada, J. Therriault, M. S. P. Kang, M. Savard, T. A. Pascoal, F. Lussier, C. Tissot, Y. Wang, A. Benedet, T. Matsudaira, T. Bunai, T. Obi, H. Tsukada, Y. Ouchi, P. Rosa-Neto, Mitochondrial complex I abnormalities is associated with tau and clinical symptoms in mild Alzheimer’s disease, Mol. Neurodegener. 16, 28 (2021).

26. W. Wang, F. Zhao, X. Ma, G. Perry, X. Zhu, Mitochondria dysfunction in the pathogenesis of Alzheimer’s disease: Recent advances, Mol. Neurodegener. 15, 1–22 (2020).

27. T. C. Südhof, Neuroligins and neurexins link synaptic function to cognitive disease., Nature 455, 903–11 (2008).

28. S. J. Finnema, N. B. Nabulsi, T. Eid, K. Detyniecki, S. Lin, M.-K. Chen, R. Dhaher, D. Matuskey, E. Baum, D. Holden, D. D. Spencer, J. Mercier, J. Hannestad, Y. Huang, R. E. Carson, Imaging synaptic density in the living human brain, Sci. Transl. Med. 8 (2016), doi:10.1126/scitranslmed.aaf6667.

29. A. P. Mecca, M. Chen, R. S. O’Dell, M. Naganawa, T. Toyonaga, T. A. Godek, J. E. Harris, H. H. Bartlett, W. Zhao, N. B. Nabulsi, B. C. Vander Wyk, P. Varma, A. F. T. Arnsten, Y. Huang, R. E. Carson, C. H. Dyck, In vivo measurement of widespread synaptic loss in Alzheimer’s disease with SV2A PET, Alzheimer’s Dement. 16, 974–982 (2020).

30. R. I. Scahill, J. M. Schott, J. M. Stevens, M. N. Rossor, N. C. Fox, Mapping the evolution of regional atrophy in Alzheimer’s disease: Unbiased analysis of fluid-registered serial MRI, Proc. Natl. Acad. Sci. U. S. A. 99, 4703–4707 (2002).

31. K. Kawamura, K. Ishiwata, H. Tajima, S.-I. Ishii, K. Matsuno, Y. Homma, M. Senda, In vivo evaluation of [11C]SA4503 as a PET ligand for mapping CNS sigma1 receptors, Nucl. Med. Biol. 27, 255–261 (2000).

32. A. Whittington, R. N. Gunn, Amyloid load: A more sensitive biomarker for amyloid imaging, J. Nucl. Med. 60, 536–540 (2019).

33. S. M. Smith, M. Jenkinson, M. W. Woolrich, C. F. Beckmann, T. E. J. Behrens, H. Johansen-Berg, P. R. Bannister, M. De Luca, I. Drobnjak, D. E. Flitney, R. K. Niazy, J. Saunders, J. Vickers, Y. Zhang, N. De Stefano, J. M. Brady, P. M. Matthews, Advances in functional and structural MR image analysis and implementation as FSL, Neuroimage 23, 208–219 (2004).

34. M. Jenkinson, P. Bannister, M. Brady, S. Smith, Improved Optimization for the Robust and Accurate Linear Registration and Motion Correction of Brain Images, Neuroimage 17, 825–841 (2002).

35. H. W. Müller-Gärtner, J. M. Links, J. L. Prince, R. N. Bryan, E. McVeigh, J. P. Leal, C. Davatzikos, J. J. Frost, Measurement of radiotracer concentration in brain gray matter using positron emission tomography: MRI-based correction for partial volume effects, J. Cereb. Blood Flow Metab. 12, 571–583 (1992).

36. R. N. Gunn, A. A. Lammertsma, S. P. Hume, V. J. Cunningham, Parametric imaging of ligand-receptor binding in PET using a simplified reference region model, Neuroimage 6, 279–287 (1997).

